# High burden of viruses and bacterial pathobionts drives heightened nasal innate immunity in children with and without SARS-CoV-2

**DOI:** 10.1101/2023.06.17.23291498

**Authors:** Timothy A. Watkins, Nagarjuna R. Cheemarla, Katrin Hänsel, Alex B. Green, Julien A.R. Amat, Richard Lozano, Sarah N. Dudgeon, Marie L. Landry, Wade L. Schulz, Ellen F. Foxman

## Abstract

Recent work indicates that heightened nasal innate immunity in children may impact SARS-CoV-2 pathogenesis. Here, we identified drivers of nasal innate immune activation in children using cytokine profiling and multiplex pathogen detection in 291 pediatric nasopharyngeal samples from the 2022 Omicron surge. Nasal viruses and bacterial pathobionts were highly prevalent, especially in younger children (81% of symptomatic and 37% asymptomatic children overall; 91% and 62% in subjects <5 yrs). For SARS-CoV-2, viral load was highest in young children, and viral load in single infections or combined viral loads in coinfections best predicted nasal CXCL10, a biomarker of the mucosal interferon response. Bacterial pathobionts correlated with high nasal IL-1β and TNF, but not CXCL10, and viral-bacterial coinfections showed a combined immunophenotype. These findings reveal virus and bacteria as drivers of heightened nasal innate immunity in children and suggest that frequent host-pathogen interactions shape responses to respiratory viruses in this age group.

## Introduction

A puzzling feature of the COVID-19 pandemic has been its lower impact in children compared to adults, prompting a search for unique features of antiviral immunity in the pediatric age group.^1^ Recently, several independent studies found heightened nasal innate immune activation in children compared to adults with SARS-CoV-2 infection and even in the absence of SARS-CoV-2 infection.^2–5^ Although there was variability among subjects, in general, nasal transcriptome patterns in children showed heightened expression of both interferon-stimulated genes (ISGs) and pro-inflammatory cytokines, and increased presence of neutrophils and pro-inflammatory monocytes in the nasal mucosa.^2–5^ Since early SARS-CoV-2 replication relies on evasion of the interferon response within its target tissue,^6, 7^ pre-activated nasal innate immunity in children has been proposed to serve as a mechanism of protection from SARS-CoV-2. However, the biological drivers of heightened nasal innate immunity in children are unknown; it is also unclear whether there are distinct patterns of heightened nasal innate immunity with different functional consequences.

While it is possible that heightened nasal innate immunity in children is due to age-intrinsic biological mechanisms, other observations suggest that these patterns may represent an interplay between microorganisms in the upper respiratory tract and the developing immune system. Children are known to have much higher rates of colonization with airway bacterial pathobionts and more frequent infections with common cold-causing respiratory viruses than adults, likely due, at least in part, to less well-developed adaptive immune defenses in this age group.^8, 9^ Recent epidemiological studies using multiplex virus detection have also found a surprisingly high rate of viral respiratory infections in asymptomatic children.^10, 11^ In some instances, asymptomatic viral and bacterial pathobiont detections have been linked to innate immune responses in the nasal mucosa.^8, 12–14^ These observations suggest that higher burdens of viral and bacterial pathobionts in the upper respiratory tract may be important drivers of the heightened nasal innate immunity seen in children. If this model is correct, virus and pathobiont loads would be expected to predict the variable nasal immunophenotypes seen within the pediatric age group.

The SARS-CoV-2 Omicron surge from December 25, 2021 – January 29, had the highest rate of reported COVID-19 cases in the U.S. to date overall across age groups, including subjects <24 years old.^15–17^ The large number of children presenting to our healthcare system for SARS-CoV-2 testing during this timeframe offered an opportunity to test the relationship between nasopharyngeal immunophenotypes and nasal viral and bacterial burden in children. Using comprehensive testing for 19 respiratory pathogens and quantitative microfluidics-based cytokine assays, we demonstrate distinct patterns of innate immune activation in pediatric subjects predicted by the type(s) and burden of microbes present in the nasopharynx. Our results indicate that the high burden of respiratory viruses and bacterial pathobionts in young children drive distinct patterns of heightened nasal innate immunity in this age group and alter mucosal responses to SARS-CoV-2.

## Results

### Children had high rates of infection with SARS-CoV-2 and other viruses in January 2022.

To better understand mucosal innate immunity during pediatric SARS-CoV-2 infection, we collected nasopharyngeal swab samples from subjects aged 0-19 years undergoing SARS-CoV-2 testing at Yale New Haven Hospital from January 11-23, 2022, during the SARS-CoV-2 Omicron surge **(Fig 1**). Samples originated in the pediatric emergency department (E.D.) from children presenting for symptoms associated with acute respiratory infection or for unrelated conditions as defined by ICD10 code, and from asymptomatic children undergoing and pre-operative (pre-op) screening for elective surgery. Among 291 samples meeting the inclusion criteria, there were 56 symptomatic SARS-CoV-2+ subjects. Of these, only 15 subjects were admitted to the hospital, including 5 admissions for infection-related symptoms, and 10 admissions attributable to comorbidities.

**Figure 1:**
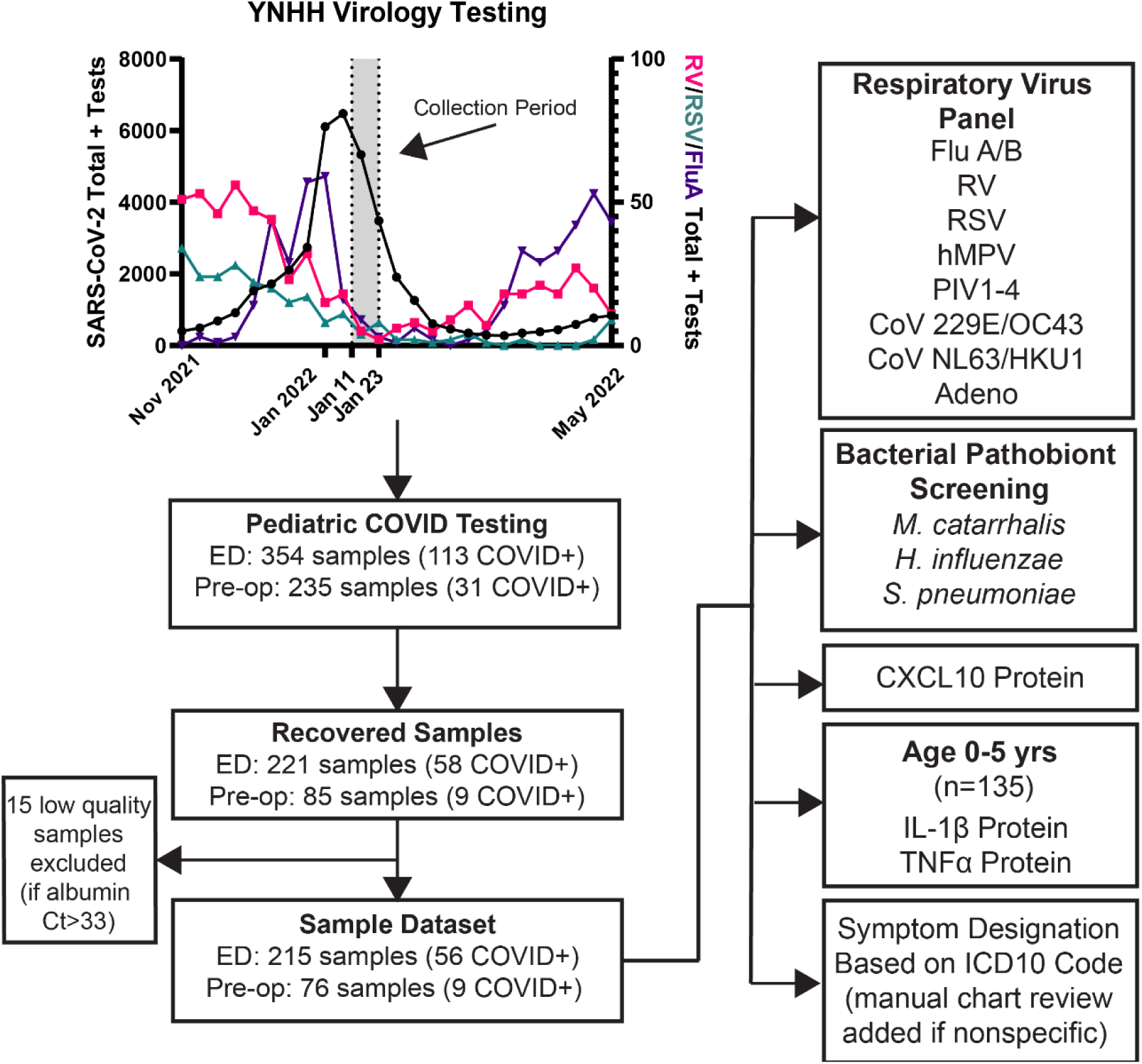
Study overview. Nasopharyngeal swab samples from subjects ages 0-19 undergoing SARS-CoV-2 testing in the YNHH pediatric E.D. or pre-operative screening were collected from January 11-23, 2022 and processed/analyzed as indicated.

RT-qPCR for 15 common respiratory viruses in 291 samples meeting the inclusion criteria showed a high prevalence of both SARS-CoV-2 and non-SARS-CoV-2 viruses in children ages 0-5 years at 54.4% (73/135), with 30.1% (40/135) SARS-CoV-2+ and 27.2% (37/135) positive for other respiratory viruses, the most prevalent being rhinovirus (RV), adenovirus (Adeno), and human metapneumovirus (hMPV) (**Fig 2A, Table S1**). Overall, the median age was significantly lower for virus-positive (2.85 years) than for virus-negative subjects (9.3 years, p<0.0001), demonstrating the association of young age with a higher prevalence of viruses in this cohort (**Table 1**).

**Figure 2:**
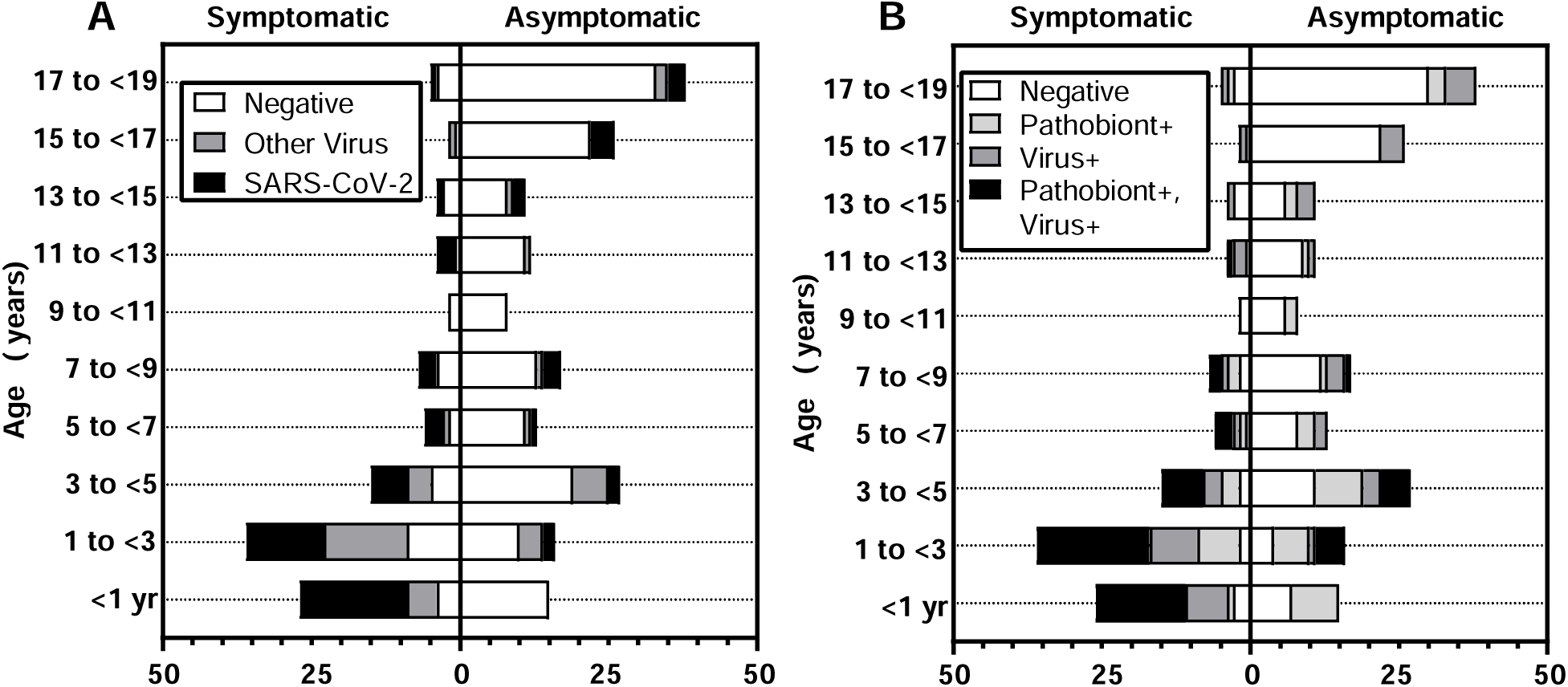
SARS-CoV-2, non-SARS-CoV-2 respiratory virus, and bacterial pathobiont test results. (A) Number of samples testing negative for 16 respiratory viruses (white) positive for non-SARS-CoV-2 respiratory virus (grey) or SARS-CoV-2 (black) by age and clinical presentation. (B) Number of samples negative for viruses and bacteria (white), positive for virus only (dark grey), pathobiont only (light grey), or virus/pathobiont double positive (black) by age and clinical presentation. See Tables S1-5 for additional information.

**Table 1:** Results of multiplex virology testing by demographics, presentation, and pathobiont status. Viral PCR results are shown with respect to biological sex, age, symptoms, and pathobiont status. P values based on Chi-square tests show relationship between number of virus positives and biological sex, symptoms, and pathobiont status. The Mann-Whitney test is used to compare number of virus positives by age. See Tables S3-5 for additional information.

|  | **Overall** | **Virus-Negative** | **Virus-Positive** | **p-value** |
| --- | --- | --- | --- | --- |
| N | 291 | 185 | 106 |  |
| **Sex** |  |  |  | **0.111** |
| Male | 144 | 85 | 59 |  |
| Female | 147 | 100 | 47 |  |
| **Median Age** | **5.8** | **9.3** | **2.85** | **<0.0001** |
| **Presentation** |  |  |  | **<0.0001** |
| Symptomatic | 108 | 35 | 73 |  |
| Asymptomatic | 183 | 150 | 33 |  |
| **Pathobiont Status** |  |  |  | **<0.0001** |
| Negative | 183 | 136 | 47 |  |
| Positive | 107 | 49 | 58 |  |

### Children had high rates of bacterial pathobiont detection and virus-pathobiont codetection

Next, we performed RT-qPCR for three common bacterial pathobionts known to colonize the upper respiratory tract and contribute to clinically significant respiratory tract coinfections: *Moraxella catarrhalis, Streptococcus pneumoniae,* and *Haemophilus influenzae.*^18–21^ All pathobionts were detected, with *M. catarrhalis* being the most prevalent (**Fig 2B, Table S1**). Median age was 2.2 years for pathobiont-positive subjects and 11.4 years for pathobiont-negative subjects (**Table S2,** p<0.0001). Among children <5 years old, 78% had viruses, pathobionts, or both, including 90% (70/77) of symptomatic subjects and 62.1% (36/58) of asymptomatic subjects. In subjects ages 5-19 years, 57% (17/30) of symptomatic and 24% (31/125) of asymptomatic subjects had viruses and/or pathobionts detected. Overall, these data reveal the high prevalence of non-SARS-CoV-2 viruses and bacterial pathobionts in the nasopharynx of children undergoing SARS-CoV-2 testing in January 2022.

### CXCL10, a biomarker of the nasal interferon response, negatively correlated with age in virus-positive subjects, but not virus-negative subjects

Previous work from our group showed that the interferon-inducible protein CXCL10 is a robust biomarker of the nasal interferon response, and that CXCL10 levels in the nasopharyngeal swab-associated viral transport media correlate directly with ISG expression by RNA-seq.^22, 23^ Therefore, to estimate activation of the mucosal interferon response in the upper respiratory tract, we measured nasopharyngeal swab-associated CXCL10 protein across all samples in the dataset using a clinical-grade microfluidics system.^24, 25^ Nasopharyngeal CXCL10 showed a significant negative correlation with age overall (**Fig 3A, dashed line**) driven exclusively by virus-positive samples (**Fig 3A, black line**). In contrast, nasopharyngeal CXCL10 showed no correlation with age in virus-negative children (**Fig 3A, grey line**). These results indicate that the heightened nasal interferon responses in young children are associated with presence of a viral infection.

**Figure 3:**
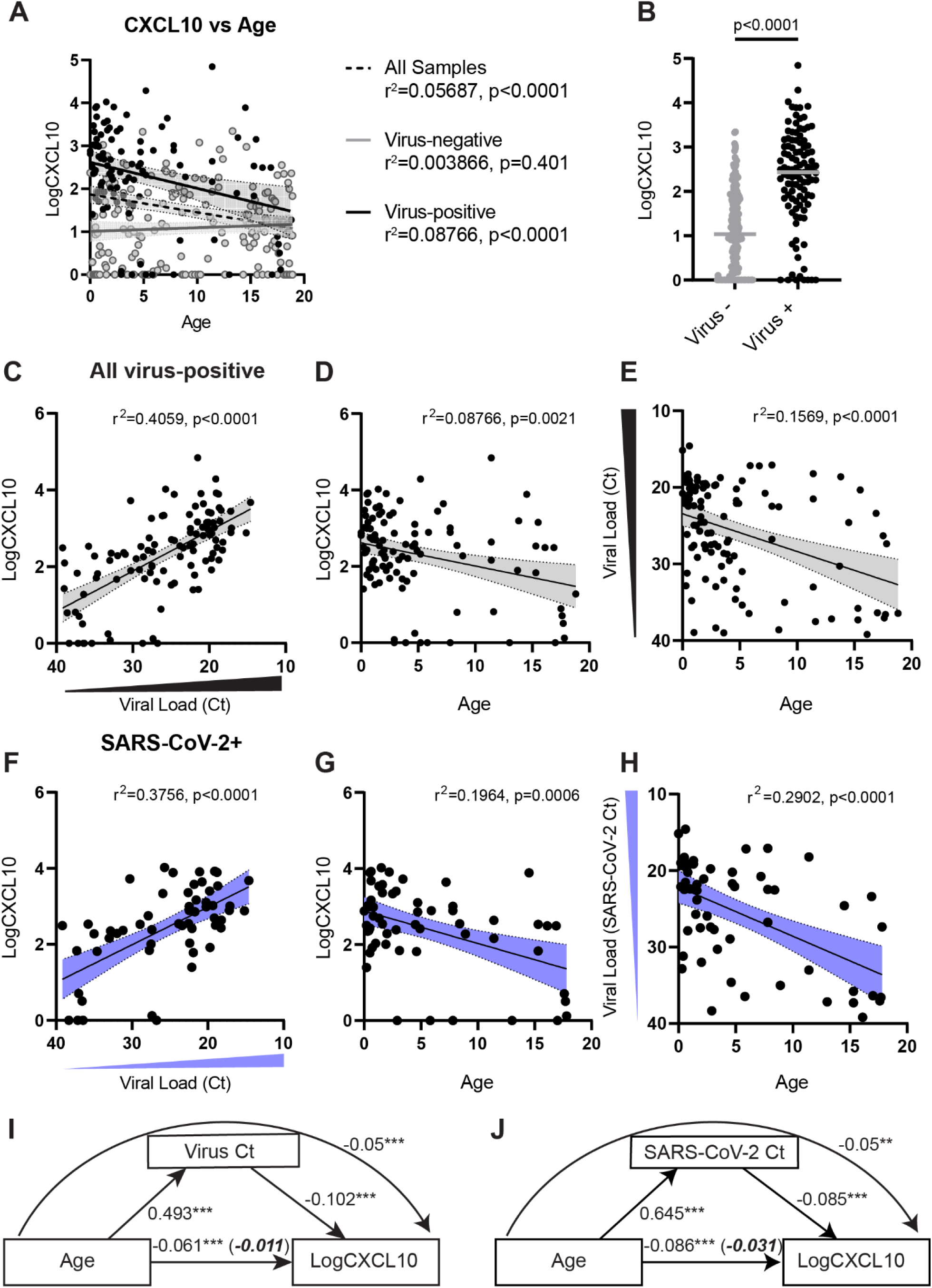
Relationship between viral load, age, and nasopharyngeal CXCL10 level in pediatric subjects ages 0-19 years. (A) Correlation between CXCL10 (Log10 pg/mL) in virus-positive subjects (black line), virus negative subjects (grey line), and all subjects (dashed line). For A and C-H, shading represents 95% CI for regression line slope, r2 and p-value for correlation are shown. (B) Nasopharyngeal CXCL10 (Log10 pg/mL) in virus-positive and virus-negative subjects of all ages (n=291). P-value by Welch’s t-test is shown. (C) CXCL10 (Log10 pg/mL) versus viral load (Ct value), all virus-positive samples. For samples with >1 virus detected, plot shows highest viral load (lowest Ct value). (D) CXCL10 (Log10 pg/mL) versus age, all virus-positive samples, (E) Viral load versus age, all virus-positive samples. For samples with >1 virus detected, plot shows highest viral load (lowest Ct value). (F) Viral load in SARS-CoV-2 single infections versus CXCL10 (Log10 pg/mL). (G) CXCL10 level (Log10 pg/mL) versus age for SARS-CoV-2 single infections. (H) Viral load versus age for SARS-CoV-2 single infections. (I) Mediation analysis results for Ct value in single infection or lowest Ct value in coinfection as a mediator between age and CXCL10. Figure shows average causal mediation effect (ACME) through viral load denoted by curved arrow (-0.05 [-0.08, -0.03 CI], p<0.001), average direct effect of age on CXCL10 (-0.0106 [-0.0476, 0.03], p=0.570), shown in italics. 95% confidence intervals for mediation analysis are derived from 1000 bootstrapped samples. (J) Mediation analysis results for SARS-CoV-2 viral load (Ct) as a mediator between age and CXCL10. Figure shows average causal mediation effect (ACME) through viral load denoted by curved arrow (-0.05 [-0.09, -0.02 CI], p=0.002), average direct effect of age on CXCL10 (-0.0307 [-0.0855, 0.03], p=0.268), shown in italics. 95% confidence intervals for mediation analysis are derived from 1000 bootstrapped samples.

### Viral load, not age, is the major driver of heightened nasal interferon response in children positive for any virus and in children positive for SARS-CoV-2

Next, we examined viral load using the cycle threshold (Ct) value from RT-qPCR testing. A lower Ct value represents a higher viral load, with each unit of change representing a 2-fold difference in viral genome copies detected. In subjects positive for any virus, amount of nasopharyngeal CXCL10 was significantly associated with virus detection (**Fig 3B**; p<0.0001) and correlated with viral load (**Fig 3C**; r^2^=0.4059, p<0.0001). We observed significant negative correlations between nasopharyngeal CXCL10 and age (r^2^=0.0876, p<0.0001) and between viral load and age (r^2^=0.1569, p<0.0001) (**Fig 3D-E**). A positive correlation between CXCL10 and viral load and negative correlations between age and CXCL10 or viral load were also observed for subjects with SARS-CoV-2 and no other virus (**Fig 3F-H**). For subjects with seasonal respiratory viruses only, viral loads also correlated significantly with CXCL10, and CXCL10 and viral load both tended to be higher with younger age, but this trend was not significant due to a limited number of samples with non-SARS-CoV-2 viruses in subjects >5 years (**Fig S1A-C**).

We applied mediation analysis with multiple regression to evaluate the effects of viral load as a mediator of the relationship between age and nasopharyngeal CXCL10 level.^26^ For virus-positive samples overall (**Fig 3I**) and for SARS-CoV-2 single infections only (**Fig 3J)** viral load fully mediated the effect on CXCL10 level. Bootstrapping analysis confirmed this result with an average causal mediation effect of -0.05 (95% CI [-0.08, -0.03], p<0.001) for all virus-positive samples and of -0.05 (95% CI [-0.09, -0.02], p=0.002) for SARS-CoV-2-positive samples. These results indicate that the higher nasal CXCL10 levels observed here in younger virus-positive children are due to a confounding effect of higher viral loads in younger children rather a direct age-intrinsic effect on the mucosal interferon response.

### Coinfection with another respiratory virus in SARS-CoV-2+ subjects is associated with a more robust mucosal antiviral response than predicted by SARS-CoV-2 viral load alone

Of the 65 SARS-CoV-2+ pediatric subjects we observed, 8 subjects had a coinfecting respiratory virus (**Table S1**). To evaluate the impact of viral coinfection on the mucosal interferon response, we compared nasal CXCL10 level to viral load for subjects with only SARS-CoV-2 detected (**Fig 4A, grey dots**) to subjects with viral coinfections (**Fig 4A, colored dots**). In subjects with SARS-CoV-2 viral coinfections, CXCL10 values were above the regression line between SARS-CoV-2 viral load and CXCL10 in single infections, suggesting more robust activation of the mucosal interferon response in coinfections (**Fig 4A**). The distance from the regression line was greatest for samples with high viral loads of coinfecting viruses including influenza A (Ct 21.5, Ct 17.2, Ct 18.6) or RSV (Ct 19.48), and low viral loads of SARS-CoV-2 (Ct 33.1, Ct 36.1, Ct 27.5, Ct 33.9). CXCL10 values fell much closer to values expected for SARS-CoV-2 single infections when subjects had high viral loads of SARS-CoV-2 and low viral loads of coinfecting viruses. These results reveal that some children diagnosed with COVID-19 based on a positive SARS-CoV-2 PCR test in fact had low viral loads of SARS-CoV-2 and much higher viral loads of other viral pathogens and suggest that the nasal interferon response during SARS-CoV-2 infection is driven by the viral load of both SARS-CoV-2 and co-infecting viruses.

**Figure 4:**
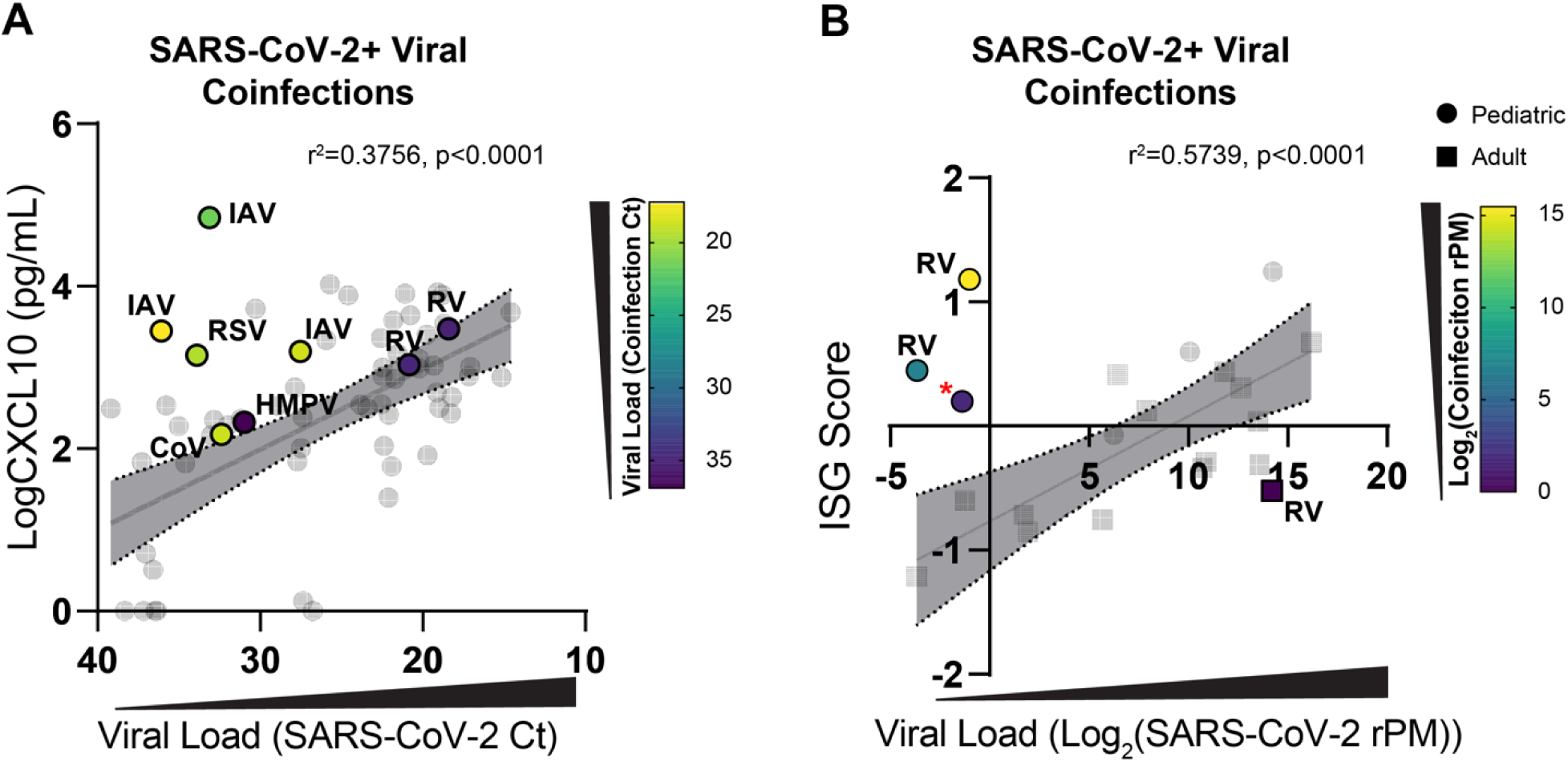
Relationship between nasopharyngeal innate immune response and viral load in SARS-CoV-2+ subjects with and without co-infecting respiratory viruses. (A) Viral load vs. nasal CXCL10 (Log10 pg/mL) concentration for pediatric SARS-CoV-2+ samples with and without viral coinfections in this study. Regression line is calculated for CXCL10 and viral load of SARS-CoV-2+ positive samples without viral coinfections (grey dots). Colored dots show viral load vs. CXCL10 SARS-CoV-2+ samples with viral coinfections. Dots are labeled with coinfecting virus and respective viral load based on the color scale. RV=rhinovirus; IAV=influenza A virus; CoV=seasonal coronavirus; HMPV=human metapneumovirus; RSV=respiratory syncytial virus. (B) Viral load vs. nasal RNA-seq ISG score for SARS-CoV-2+ samples with and without viral coinfections from children (circles) and adults (squares), GSE17274 dataset. Regression line is calculated for ISG score and viral load for SARS-CoV-2+ positive samples without viral coinfections (grey symbols). Colored symbols show samples with viral coinfections. The red asterisk denotes a patient with reads from attenuated measles vaccine virus. See Table S6 for additional information.

To further probe whether coinfections impact nasal interferon responses during SARS-CoV-2 infection, we analyzed RNA-seq data from a previous study showing heightened nasal innate immunity in SARS-CoV-2+ children (GSE172274), using ISG expression to estimate the nasal interferon response and metatranscriptomics to identify viral reads as previously described ^2, 24, 27–29^. Coinfecting viruses were identified in 3/6 children, all <5 years old, including two subjects with high rhinovirus viral loads (14575 reads per million (rPM) and 1036 rPM) and SARS-CoV-2 reads near or below the limit of detection from RNA-seq (0.4902 rPM and 0 rPM). One sample contained reads from the vaccine strain of measles virus consistent with age of the subject being typical for the first MMR vaccination, and case reports that vaccine measles virus can be found nasopharynx 7-15 days post-vaccination.^30–32^ In contrast, only one viral coinfection was identified among the 15 adults with nasal RNA-seq data, with low rhinovirus viral load (52 rPM) and high viral load of SARS-CoV-2 (18821.5 rPM). ISG score directly correlated with viral load in subjects with SARS-CoV-2 and no other virus detected; however, the three children with non-SARS-CoV-2 viral coinfections had higher ISG scores than expected based on the relationship between SARS-CoV-2 viral load and ISG score in single infections (**Fig 4B**). While the number of subjects is limited, these findings support the idea that coinfecting viruses drive an augmented mucosal interferon response during SARS-CoV-2 infection, and that more frequent viral coinfections in children may contribute to the previously described heightened nasal immune responses in SARS-CoV-2-positive children compared to adults.

### Bacterial pathobionts alter the nasal mucosal immunophenotype

We next sought to explore whether bacterial pathobiont detection correlates with changes in nasal mucosal innate immune activation. For this analysis, we focused on children under the age of 5 years in whom pathobionts were most prevalent.

To select nasopharyngeal biomarkers that might capture pathobiont-induced mucosal innate immune responses, we examined differentially-enriched transcripts and cytokines from a previous study in which we identified nasopharyngeal samples from rhinovirus-infected children and adults, about half of whom showed high loads of bacterial pathobionts based on bacterial reads in RNA-seq data.^24^ Pathways enriched in rhinovirus-positive pathobiont-high samples compared to rhinovirus-positive pathobiont-low samples indicated leukocyte recruitment, myeloid cell activation, NFkB-signaling, and interferon signaling (**Fig S2A**). Among the top 10 cytokine regulators of DEGs, 4 were also highly differentially expressed at the mRNA level: TNF, IL-1β, IL-6, and IL-1α (**Fig S2B**). Multiplex testing for 71 cytokines in these samples revealed that TNF and IL-1β were also among the top 10 differentially expressed cytokines in the nasopharynx at the protein level (**Fig S2C**). IL-1β and TNF were also shown to be associated with colonization of *M. catarrhalis* and *H. influenzae* in the airway in a prior study of 662 asymptomatic infants, suggesting that bacterial pathobionts also induce these cytokines in the nasal mucosa without rhinovirus infection, although the presence of viruses was not assessed in the prior study.^14^

To explore TNF and IL-1β as biomarkers of the mucosal innate immune response induced by bacterial pathobionts, we evaluated relationships between age, pathobionts, and nasopharyngeal protein levels of IL-1β and TNF in children <5 years. IL-1β was significantly elevated in pathobiont-only detection and was similarly elevated in virus/pathobiont codetection (**Fig 5A**). TNF was modestly but significantly elevated in pathobiont-only samples (p=0.0224) and enhanced by virus/pathobiont codetection (p<0.0001; **Fig 5B**). Pathobiont load in all pathobiont-positive samples as well as *M. catarrhalis*-only samples correlated directly with IL-1β level, further suggesting pathobiont load is a driver of IL-1β level (r^2^=0.2843, p<0.0001; r^2^=0.2562, p<0.0001) (**Fig 5C-D**). TNF level correlated significantly with pathobiont load in both virus-positive and virus-negative samples, and the slope of virus-positive samples trended slightly higher, but the difference was not significant (p=0.1105) (**Fig 5E**). These data suggest that nasopharyngeal IL-1β and TNF reflect a pathobiont-induced mucosal inflammatory response that occurs independently of, but may be enhanced by, viral coinfection.

**Figure 5:**
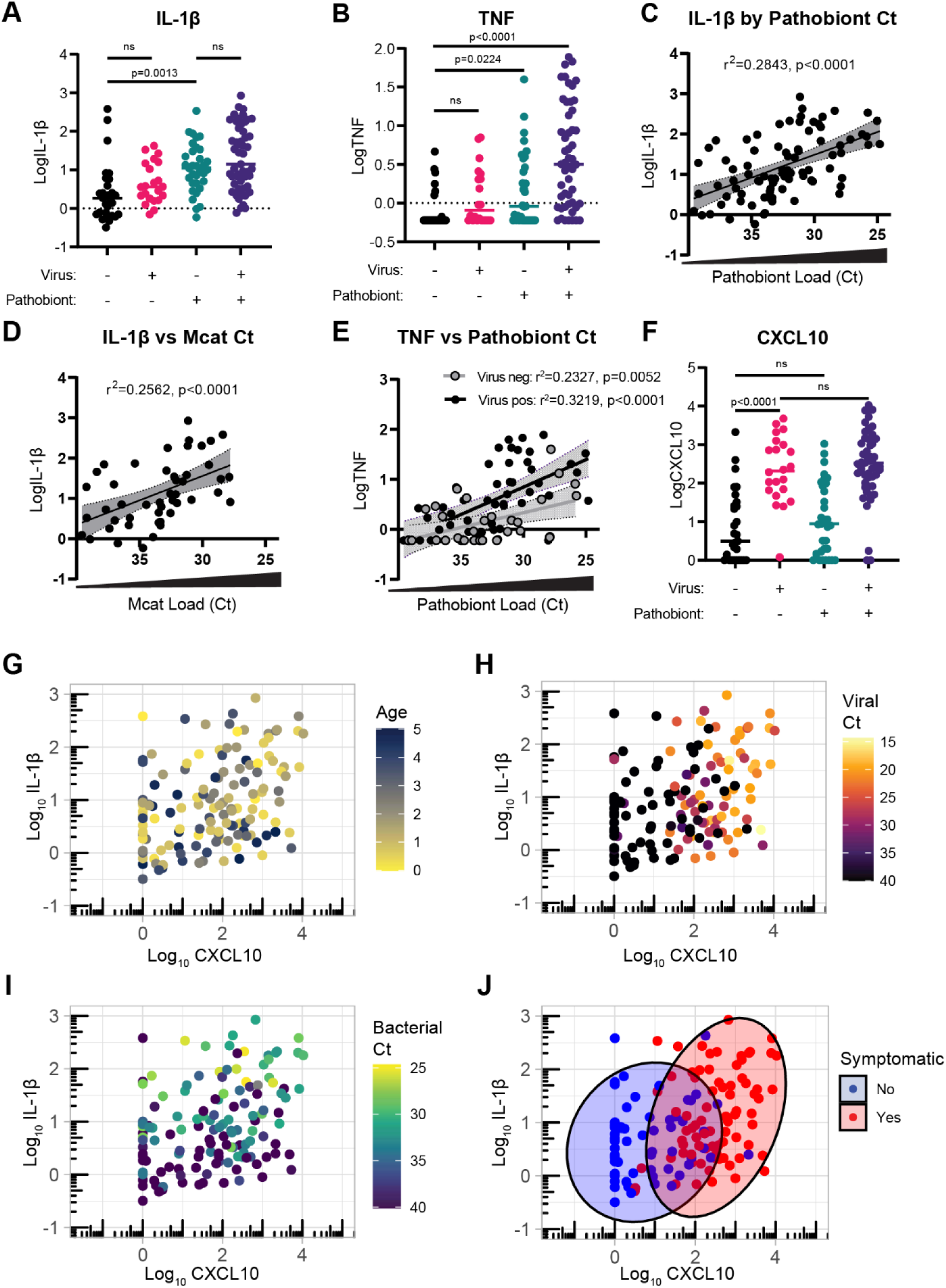
Immunophenotyping of nasal mucosal innate response in children <5 years. (A) Nasal IL-1β protein concentration (Log_10_pg/mL) in 135 nasopharyngeal samples from children under 5 years with or without viruses or pathobionts detected. P-values based on Welch’s ANOVA test with Dunnett’s T3 multiple comparisons tests are shown. ns=not significant. (B) Nasal TNF protein concentration (Log_10_pg/mL) in virus-/pathobiont-, virus-/pathobiont+, virus+/pathobiont-, and virus+/pathobiont+ samples. P-values for groups with significant differences based on Kruskal-Wallis test with Dunn’s multiple comparisons tests are shown. Ns=not significant (C) IL-1β (Log_10_pg/mL) vs. pathobiont load (Ct value) in pathobiont-positive samples. For samples with >1 pathobiont detection, lowest Ct value is plotted. Simple linear regression with 95% confidence bands is plotted for D-F. (D) IL-1β (Log_10_pg/mL) vs. *M. catarrhalis* load (Ct value) in samples positive for *M. catarrhalis* only and no other pathobionts. (E) TNF (Log_10_pg/mL) vs. pathobiont load (Ct value). Points show virus-negative samples (open circles), virus-positive samples (black circles). Linear regression for virus-negative samples (grey line) and virus-positive samples (black line) are shown. P-value for difference between slopes is shown (p=0.1055, n.s.). (F) CXCL10 protein concentration (Log_10_pg/mL) in virus-/pathobiont-, virus-/pathobiont+, virus+/pathobiont-, and virus+/pathobiont+ samples. P-values based on Welch’s ANOVA test with Dunnett’s T3 multiple comparisons tests are shown. ns=not significant. Statistical comparisons are made using the Welch’s ANOVA test with Dunnett’s T3 multiple comparisons tests. (G) –(J) Log_10_CXCL10 vs. Log_10_IL1-β for subjects <5 years of age (n=135). Color scaled by age (G), viral Ct (H), bacterial Ct (I), and symptom status (J). For (J), ellipses cover 80% of points in each group. See figures S3-5 for additional information.

We also explored the impact of pathobiont detection on nasal CXCL10 in the same four groups. Consistent with the sample set as a whole (**Fig 2**), in children <5 years, we observed significant elevation in nasal CXCL10 in virus-positive samples compared to virus-and pathobiont-negative controls regardless of pathobiont detection, but no CXCL10 elevation in pathobiont-positive, virus-negative subjects. (**Fig 5F**). When limiting the analysis to asymptomatic children, nasal CXCL10 was also elevated in virus-positive compared to virus-negative subjects, although asymptomatic subjects had lower viral loads and CXCL10 levels than symptomatic subjects (**Fig S3A-B**). Together, these data provide evidence that a robust mucosal interferon response, as indicated by nasopharyngeal CXCL10 elevation, is triggered by viral infection but not bacterial pathobionts.

Since pathobionts are associated with clinically significant secondary infections following viral infection, we also explored the relationship between pathobiont detection and viral load. Among children <5 years of age, neither viral load nor pathobiont load was significantly different in viral-bacterial codetections compared to single detections, and pathobiont loads and viral loads did not correlate with each other in codetections (**Fig S3C-E**). We also did not observe a significant change of nasal immunophenotypes defined by IL-1β, TNF, and CXCL10 levels attributed to patient sex in this age group (**Fig S4A-C**). Together, these findings indicate that both viruses and bacterial pathobionts promote heightened nasal innate immunity in young children, and that viruses and pathobionts drive distinct patterns of heightened nasal innate immunity.

### Biomarkers demonstrate heightened nasal innate immunity in children <5 years old with distinct patterns related to viral and bacterial pathobiont burden

To explore associations of age, viruses, pathobionts, and symptoms with nasal immunophenotypes in children <5 years of age, we generated scatterplots based on concentrations of CXCL10 and IL-1β, the nasopharyngeal biomarkers best associated with viral infection and bacterial pathobionts respectively. Within the 0-5 years age group, age did not appear to correlate with cytokine responses (**Fig 5G**). In contrast, plots highlight a strong association between viral load and nasopharyngeal CXCL10 and between bacterial load and nasopharyngeal IL-1β (**Fig 5H-I**). Children presenting to the E.D. with symptoms of acute respiratory infection separated into a CXCL10-high and IL-1β low-to-high phenotype, indicating that viral infection was highly associated with symptomatic presentation (**Fig 5J**). Among virus-positive subjects <5 years old, symptomatic subjects and asymptomatic subjects were equally likely to be pathobiont-positive suggesting that pathobiont detection was not a strong influence on symptomatic presentation (**Table S2**). However, virus-positive, pathobiont-positive subjects did have an enhanced mucosal inflammatory response as indicated by trend toward higher nasal TNF in subjects with viral/pathobiont codetection compared to those with virus or pathobiont only (**Fig 5B, E**). Taken together, these analyses show that viruses and nasal bacterial pathobionts are associated with distinct patterns of heightened mucosal innate immunity, alone and in combination, compared to age-matched virus-negative, bacteria-negative controls.

## Discussion

Innate immune defenses at the site of infection are critically important in limiting susceptibility to respiratory viruses, particularly in the case of emerging viruses such as SARS-CoV-2 when there is no prior adaptive immunity. Here we show a clear relationship between viral presence and abundance in the upper respiratory tract and activation of the interferon response, a potent antiviral defense pathway. We also show a link between bacterial pathobiont burden and activation of pro-inflammatory nasal mucosal responses. Together with prior studies, these results point to a high burden of respiratory viruses and bacterial pathobionts in children and link these mucosal pathogens with heightened nasal innate immunity. Our results also highlight examples of viral-viral and viral-bacterial coinfections which illustrate that the host response to one microbe influences the mucosal response to another in the airway niche.

A key feature of our study was performing PCR testing targeting 15 seasonal respiratory viruses and three common bacterial pathobionts in addition to SARS-CoV-2, including samples from symptomatic and asymptomatic subjects. RT-qPCR is a sensitive and specific assay for respiratory viruses in clinical samples. In prior studies of pediatric nasal innate immunity, samples were tested by RT-qPCR for SARS-CoV-2 but not for other respiratory viruses.^2–5^ With RT-qPCR testing for 16 viruses, we found a high burden of seasonal respiratory viruses in addition to SARS-CoV-2 during the January 2022 Omicron surge that were not diagnosed at the time of SARS-CoV-2 testing. We were also able to examine the quantitative relationship between viral loads and mucosal cytokine responses using sensitive microfluidics-based immunoassays with high accuracy and large dynamic range.^25^

Comparing nasal cytokine responses to virus detections and loads in individual samples revealed viral load as the proximal driver of the nasal interferon response in children. First, in SARS-CoV-2 single infections or with other respiratory viruses, younger children had heightened nasal interferon responses, but this was largely due to higher viral loads in younger children. Examining non-COVID viruses in SARS-CoV2+ subjects revealed coinfections as an explanation for the disparity between SARS-CoV-2 viral load and nasal interferon response in prior studies. Interestingly, we also found a higher-than-expected nasal interferon response in a SARS-CoV-2+ young child with nasopharyngeal co-detection of measles vaccine strain virus, suggesting that live attenuated vaccines may also impact heightened nasal innate immunity in children. Intuitively, the finding that the upper respiratory tract interferon responses correlate with viral load fits with the known mechanism of interferon and ISG induction, in which viral RNA recognition by innate immune sensors initiates the signaling cascade leading the interferon response.^33^

There are several possible reasons for higher prevalence and load of respiratory viruses among the youngest children in this data set. First, we considered biases in the sample set; for example, younger children were more likely to present to the ED with acute respiratory illness, whereas older children were more likely to present for other reasons. However, even among asymptomatic subjects, the prevalence of both SARS-CoV-2 and seasonal respiratory viruses was the highest in younger children, suggesting greater susceptibility. Also, young children have been shown to have higher prevalence of seasonal respiratory viruses than older children and adults with unbiased sampling. For example, in a year-long study of 26 families which included weekly nasal sampling agnostic of symptoms, respiratory viruses were detected on average 26 weeks per year in children under 5 with decreasing detection rates for older age groups.^10^ Public health interventions drastically reduced circulation of many respiratory viruses during the first year of the COVID-19 pandemic; however, there is evidence for increased circulation of rhinoviruses and enteroviruses by June 2020, for adenoviruses, seasonal coronaviruses, and parainfluenza by early 2021, and other viruses later in 2021.^34^ Our results demonstrate high circulation of diverse non-SARS-CoV-2 viruses in young children by January 2022.

The higher susceptibility of young children to seasonal respiratory viruses is generally attributed to less adaptive immunity due to fewer prior exposures.^35^ For SARS-CoV-2, a novel virus which is thought to have first become widespread in children during the Omicron surge, we were initially surprised to also observe higher prevalence and viral loads in the youngest children among our study subjects aged 0-19 yrs. While greater adaptive immunity in older children due to more prior exposure is a possibility, vaccination may have also played a role, since vaccines were only available to children >5 years old in the U.S. in January 2022.^36, 37^ It is also possible that infants and young children were more likely to present for healthcare than older children with similar symptoms, and symptomatic subjects had higher viral loads. Other features of upper respiratory tract biology in early life could also play a role. For example, there is some evidence that unique features of the pediatric nasal mucosa promote replication of some respiratory viruses based on ex vivo culture.^38^

Relevant to understanding the roles of cell intrinsic vs environmental factors in heightened innate immunity in children, a recent study by Maughan et al. compared airway mucosa of children and adults in vivo and ex vivo, using both laser capture-microdissection of biopsies, airway basal cells sorted directly from biopsies, and proliferating primary basal epithelial cells in culture.^39^ RNA sequencing from biopsies and sorted basal cells, which captured the transcriptome at the time of sampling, recapitulated previous findings from patient samples, showing that interferon response pathways are elevated in children compared to adults. However, basal cells cultured in vitro from the same subjects showed a trend toward lower innate immune activation in epithelial cells derived from children. This study is consistent with our findings indicating that viruses and bacteria found in the nasal mucosa in vivo, rather than cell-intrinsic factors, drive heightened respiratory mucosal innate immunity in children.

In addition to heightened interferon responses, prior studies using scRNA-seq of patient samples showed that the pediatric nasal mucosal was enriched for leukocytes, particularly neutrophils and pro-inflammatory monocytes.^3, 4^ We observed similar myeloid-rich leukocyte signatures in nasopharyngeal RNA-seq data from rhinovirus-infected subjects with high levels of bacterial pathobionts compared to pathobiont-low, rhinovirus-infected subjects, associated with increase in nasal IL-1β and TNF proteins. Analysis of samples in this study confirmed nasal IL-1β and TNF as biomarkers of pathobiont-associated inflammation in both virus-negative and virus-positive subjects. Pathobionts alone did not induce nasal interferon responses, as indicated by no change in the CXCL10 level, but were associated with a synergistic increase in TNF. Together, these observations show that pathobionts promote local mucosal inflammatory responses and may enhance antiviral responses upon viral coinfection.

While our study shows a high frequency of pathobionts in young children, it is still unclear if pathobionts or the inflammatory responses they induce are protective or detrimental in COVID-19. For seasonal respiratory viruses, bacteria in the genera *Moraxella, Haemophilus,* and *Streptococcus* increase the incidence of infection, disease severity during infection, risk of lower respiratory tract infection, and later asthma development.^40–43^ This evidence is particularly strong for RSV, influenza, and rhinoviruses.^44, 45^ We found no difference in SARS-CoV-2+ viral loads in age-matched pathobiont-positive and pathobiont-negative subjects under the age of 5 years, but SARS-CoV-2+ samples in this age group were almost all from symptomatic subjects with similar high viral loads. Evaluating this question with unbiased sampling, including longitudinal studies on children with and without pathobionts at baseline, will provide more insight into how pathobionts influence SARS-CoV-2 susceptibility and outcomes.

Our study also demonstrated high prevalence of respiratory viruses in children and induction of mucosal interferon responses concomitant with viral load. Together with the prior literature, our data support a model in which children are highly susceptible to respiratory viruses but also that frequent host-virus interactions have a net effect of limiting closely spaced infections by activating mucosal antiviral defenses. Recent experimental evidence for interferon-mediated interference among respiratory viruses supports this idea. For example, recent work from our group and others shows that prior infection with rhinovirus and other seasonal respiratory viruses can induce a robust interferon response that reduces replication of SARS-CoV-2 or influenza viruses in simultaneous or sequential infections.^23, 46–50^ Also compelling is a recent vaccination study by Costa-Martins et. al., which showed that among asymptomatic children ages 2-5 receiving the live attenuated influenza vaccine, 41% had seasonal respiratory viruses detected, virus detection was associated with a nasal ISG signature, and nasal ISG expression prior to vaccination correlated with reduced replication of vaccine viruses.^11^ While the outcome of viral coinfections depends on many factors including the viruses involved, relative timing, and host susceptibility, we propose that when the first infection is well-controlled by the mucosal interferon response, the effect is likely to be protective, such as in the setting of asymptomatic or resolving viral infections that are common in young children.

In sum, this qualitative and quantitative analysis of respiratory viruses, bacterial pathobionts, and cytokine biomarkers reveals viruses and bacterial pathobionts as major drivers of heightened nasal innate immunity in children and compels further study of how seasonal respiratory viruses and bacterial pathobionts impact SARS-CoV-2 infection in children, including interactions occurring in asymptomatic subjects.

### Limitations of the study

Here we measured a targeted panel of cytokine biomarkers, viruses and bacteria in a large sample set, complementing prior work which used detailed immunophenotyping by RNA-seq and single cell sequencing albeit in smaller sample sets. We focused on 19 respiratory microbes, but other less common viruses or nasopharyngeal pathobionts could also contribute to nasal innate immune activation in children. Future studies pairing detailed immunophenotyping with sensitive pathogen detection methods and clinical data will provide further insights into the range of nasal immunophenotypes in children and their drivers. Additionally, this analysis captured only a small number of viral coinfections with SARS-CoV-2 which suggested that viral coinfections augment the interferon response to SARS-CoV-2, but further studies are required for confirmation. Finally, we studied a cross-section of samples from a single time point. Future longitudinal studies tracking how nasal microbes and immunophenotypes in children impact SARS-CoV-2 infection in will provide further insights into host-pathogen interactions and the consequences of heightened mucosal innate immunity in children.

### Resource Availability

This study did not generate new unique reagents.

### Lead Contact

Further information and requests for resources and reagents should be directed to and will be fulfilled by the lead contact, Ellen Foxman.

### Data and code availability

All data produced in the present work are contained in the manuscript. Extended data analyses, original code for mediation analysis and data visualizations, and extended demographics data will be deposited in Mendeley Data at the DOI 10.17632/g8ckr9zxbx.1 and will be available at the time of peer-reviewed publication.

## Experimental procedures

### Nasopharyngeal swab collection and inclusion criteria

In this study, we collected a total of 306 residual nasopharyngeal swab samples from patients presenting to the pediatric emergency department between January 11-23, 2022 and were screened for SARS-CoV-2. 15 samples were excluded from further analysis due to low Ct for the internal control (albumin, Ct>33), leaving 291 samples for analysis. The study protocol was reviewed and approved by the Yale Human Investigation Committee (protocol #2000027656) and was determined to not require specific patient consent.

### Chart review for symptom designation and admission comorbidities

To assign symptoms, we extracted, de-identified ICD-10 codes and supplemented with manual chart review. Patients were considered “symptomatic” if the ICD-10 code associated with presentation to the ED are common in respiratory infections. For patients with acute, symptomatic presentations related to respiratory infections, ICD-10 codes alone were used to ascribe clinical syndromic categories. Where ICD-10 codes for acute, symptomatic presentations were ambiguous (e.g., “Emergency use of U07.1 | COVID-19”), or for clinical samples which were obtained in the preoperative setting, manual chart review by a clinical reviewer blinded to bacterial, viral, and immune biomarker status was conducted for determination of clinical syndromic category. Detailed examination of patients positive for COVID-19 testing and admitted to the inpatient setting were characterized via manual chart review to determine presenting symptoms and comorbidities. In this latter instance, the clinician reviewer was blinded to bacterial status, common respiratory viral status, and biomarker status, but not COVID-19 status.

### Cytokine measurements

Viral transport media was thawed on ice before measurement of cytokines and subsequently aliquoted and stored at −80°C for use in other experiments. CXCL10, IL-1β, and TNF proteins were measured using the Ella automated microfluidics system (Protein Simple, San Jose, USA).

### Clinical Virology Testing

For testing by the YNHH Clinical Virology Laboratory, nasopharyngeal swabs were placed in viral transport media (BD Universal Viral Transport Medium) immediately upon collection for clinical SARS-COV-2 testing. Viral transport media associated with nasopharyngeal swasb was aliquoted and stored at −80°C for further analyses within three days of clinical testing. For virology testing, 200 μL of VTM were used for total nucleic acid extraction using the NUCLISENS easyMAG platform (BioMérieux, France). Extracted nucleic acid was tested for a 15-virus panel as described previously.^24^ SARS-CoV-2 testing was completed using four clinical testing platforms. When the clinical testing platform did not result in a Ct value for SARS-CoV-2 or the value was unable to be retrieved from medical records, we performed in-lab SARS-CoV-2 qPCR testing as follows: 140 μL of viral transport media was used for viral RNA extraction using the using the QIAamp Viral RNA Kit (QIAGEN, Venlo, ND). 5 μL of viral RNA eluate was then used to generate cDNA using the iScript cDNA synthesis kit (Bio-Rad, CA, USA). 2 μL of cDNA was then used to test for SARS-CoV-2 using the 2019 CDC SARS-CoV-2 N1 assay (IDT, IA, USA)^51^. If the SARS-CoV-2 E gene Ct value was measured in either the Roche cobas or Cepheid Xpress Xpert test, this value was used for analysis. If the SARS-CoV-2 E gene measurement was not retrieved from the medical record or was not tested on either the Roche or Cepheid platforms, we converted Ct values from other platforms and genes to the SARS-CoV-2 E gene using interpolation of the linear regression between samples with known E genes and other SARS-CoV-2 genes reported. This approach applied to the Roche cobas SARS-CoV-2 ORF1ab test, the Cepheid SARS-CoV-2 N2 test, and the CDC SARS-CoV-2 N1 test. For SARS-CoV-2 and all other respiratory viruses, a Ct value of 40 or above is considered negative.

### Pathobiont RT-qPCR

All samples of sufficient quality and sample amount were tested for *Moraxella catarrhalis, Haemophilus influenzae, and Streptococcus pneumoniae* (n=290). One sample present in the multiplex virus-tested dataset was excluded from pathobiont RT-qPCR due to low sample amount and is not included in immunophenotyping analysis, leaving a sample set of 290. For measurement of pathobionts, 2 μL of total nucleic acid extraction from clinical testing was used for each pathobiont RT-qPCR assay. The AgPath-ID One-Step RT-PCR master mix (Applied Biosystems, MA, USA) was used with commercially available Taqman qPCR assays designed for each pathobiont (ThermoFisher, MA, USA). Thermocycler conditions were set according to the manufacturer’s instructions.

### Quantification and Statistical Analysis

GraphPad Prism (version 9.5.1; GraphPad Software, San Diego, CA, USA) was used for simple linear regression analyses, Welch’s ANOVA tests with Dunnett’s T3 multiple comparisons tests, Kruskal-Wallis tests with Dunn’s multiple comparisons tests, and unpaired t-tests as specified in figure legends. For mediation analysis, we considered two different viral load categories. First, for all virus-positive individuals (n = 106), we considered the Ct value of the infecting virus for single infections and the minimum Ct value (i.e., highest viral load) virus for individuals identified with coinfections. Second, we focused on individuals with a SARS-CoV-2 single infection (n=57) where only the Ct value of the SARS-CoV-2 test was considered. We followed Baron and Kenny’s causal step approach using linear modelling of the effect.^26^ Pre-requisites for indicating the appropriateness of the mediation analysis were confirmed by testing for a significant direct effect from the independent variable (IV; i.e., age) towards the dependent variable (DV; i.e., log_10_-transformed CXCL10) and a significant effect of the IV on the mediator (i.e., viral Ct). The mediation effect is then tested by considering the combined effect of the mediator and IV on the DV. In case the significant effect of the IV on the DV is reduced by the inclusion of the mediator, we can positively confirm the mediation. Lastly, casual mediation effects were computed for 1000 bootstrapped samples and the 95% confidence-interval is reported. The analysis was performed using RStudio version 2022.12.0.353 using the stats package 4.1.2 for the linear modelling and mediation 4.5.0 for building the final mediation model and extracting information on the causal mediation effects.^52, 53^

### Data Visualization

GraphPad Prism (version 9.5.1; GraphPad Software, San Diego, CA, USA) was used to generate graphs for the majority of main and supplemental figures. For figure 5G-H, RStudio (version 4.2.2) software was used.^53^ Data curation and preparation was conducted through use of tidyverse (version 2.0.0) and lubridate (version 1.9.2) R packages, while visualization was assisted by the ggplot2 (version 3.4.1) package.^54–56^ For figure S2, Ingenuity Pathway Analysis software (QIAGEN Digital Insights) and Qlucore Omics Explorer (Qlucore) were used to generate graphs and heatmaps. All figures were edited and arranged in Adobe Illustrator (Adobe Inc, Mountain View, CA, USA).

### RNA-seq Analysis of GSE172274

RNA-seq analysis of samples deposited in the NIH Gene Expression Omnibus (GEO) database under the accession number GSE172274 was carried out using Partek Flow software (version 10.0) for human transcripts.^2^ Samples were aligned to the GRCh38 human genome using Bowtie 2 and quantified using Ensembl Transcripts Release 104.^27, 28, 57^ Counts were normalized as counts per million (CPM) plus 0.0001 for downstream log-scaling analysis. Analysis focused on 50 genes found to be upregulated and differentially expressed in nasal epithelial cells during the antiviral response from a previous dataset.^22^ Normalized counts were converted to Z-scores by subtracting the mean expression of each gene across all sample from each sample count and dividing by the standard deviation of the gene’s expression across all samples. Finally, the ISG score was calculated by averaging the Z-scores for the 50 listed genes for each sample. Metatranscriptomics analysis of samples in GSE172274 were analyzed using the CZ ID metatranscriptomics pipeline as previously described.^24, 29^ Non-SARS-CoV-2 respiratory virus reads were considered positive if above 1 read per million (rPM). All samples in this dataset tested positive for SARS-CoV-2 by clinical testing as described previously.^2^

### Pathobiont Marker Selection and Pathway Analysis

RNA-seq analysis was performed on datasets available on the NIH Database of Genotypes and Phenotypes (dbGaP) under accession codes phs002442.v1.p1 and phs002433.v1.p1 as previously described.^23, 24^ Ingenuity pathway analysis (IPA) was carried out using differentially expressed genes (DEGs) between rhinovirus-positive/pathobiont-high and rhinovirus-positive/pathobiont-low samples to identify genes expressed in virus/pathobiont codetection.

Analysis of upstream regulators of DEGs listed in results was filtered on cytokines only. For multiplex protein analysis, Qlucore Omics Explorer was used to identify differentially expressed cytokines. Cytokines with <1 pg/mL expression across all samples were excluded, and remaining cytokines were log_2_-scaled. Finally, two-group comparison between rhinovirus-positive/pathobiont-high and rhinovirus-positive/pathobiont-low samples was used to narrow down to the top 10 differentially expressed cytokines between these groups and are listed in order of significance of the q-value statistic. Only samples that are virus-negative/pathobiont-low, rhinovirus-positive/pathobiont-low and rhinovirus-positive/pathobiont-high are shown.

## Acknowledgments

This study was funded by the National Institutes of Health (T32AI055403 received by TAW), Fast Grants for COVID-19 research from the Mercatus Center (George Mason University, Fairfax, VA, USA; received by EFF), the Rita Allen Foundation (received by EFF) and the Gruber Foundation (received by TAW). We thank Bao Wang, Valia Mihaylova, and Hui Jing Lim for helpful discussions; Rolando Garcia-Milian for training and assistance with Partek Flow software, Qlucore Omics Explorer, and Ingenuity Pathway Analysis; the Yale New Haven Hospital clinical virology laboratory for valuable assistance and support.

## Contributions

Conceptualization: EFF. Methodology: TAW, NRC, KH, ABG, WLS, EFF. Software: KH, JARA, SND, WLS. Validation: TW, WLS, EFF. Formal Analysis: TAW, KH, JARA, SND. Investigation: TAW, NRC, ABG, RL, EFF. Resources: MLL (nasopharyngeal swabs). Data Curation: TAW, NRC, KH, JARA, and ABG. Writing – Original Draft: TAW and EFF. Writing – Review & Editing: all authors. Visualization: TAW, KH, and JARA. Supervision and Funding Acquisition: EFF.

