## Supplementary material for "High burden of viruses and bacterial pathobionts drives heightened nasal innate immunity in children with and without SARS-CoV-2": Data supplement

**Supplemental Figures and Tables**

**Figures**

**Figure S1 (related to Figure 3):** Non-SARS-CoV-2 viruses and correlations with CXCL10 and age.

**Figure S2 (related to Figure 5):** Cytokines and pathways differentially expressed between rhinovirus-positive/pathobiont high vs. rhinovirus-positive/pathobiont low nasopharyngeal samples.

**Figure S3 (related to Figure 5):** Nasopharyngeal CXCL10 levels and viral loads in symptomatic and asymptomatic virus-positive and virus-negative subjects.

**Figure S4 (related to Figure 5):** Mucosal cytokines in relation to virus and/or pathobiont positivity and biological sex.

**Supplemental Tables**

**Table S1 (related to Figure 2 and Table 1):** Number of samples testing positive for respiratory viruses and pathobionts.

**Table S2 (related to Figure 2, Table 1):** Overview of pathobiont test results by demographics, presentation, and viral status.

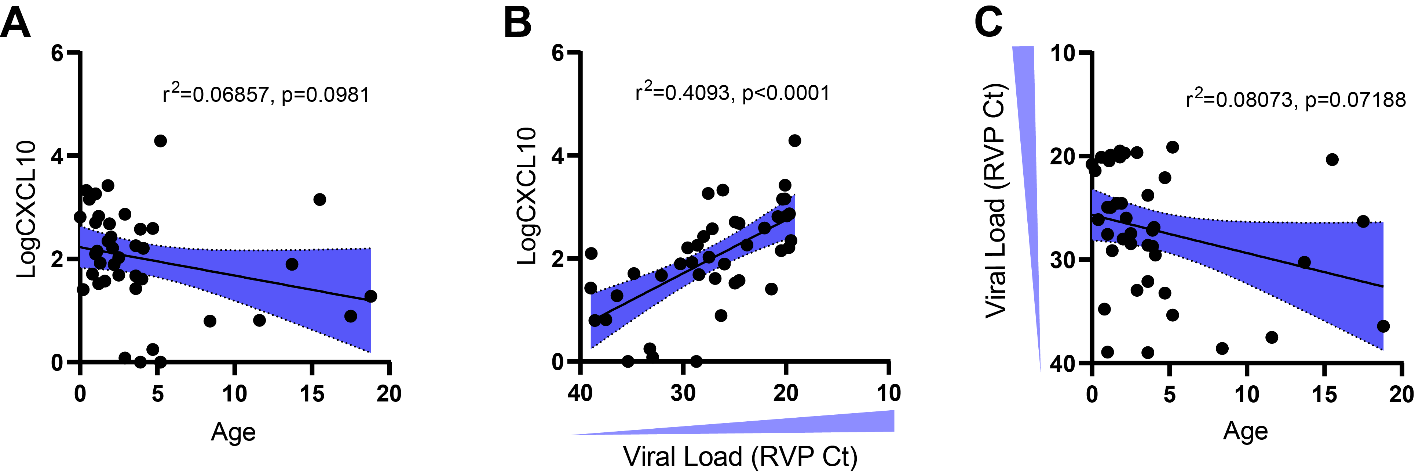

**Figure S1 (related to Fig 2): Non-SARS-CoV-2 viruses and correlations with CXCL10 and age.**

(A) For non-SARS-CoV-2 respiratory viruses (n=41), CXCL10 (Log_10_pg/ml) is plotted against age.
(B) Viral load of non-SARS-CoV-2 viruses vs. CXCL10 (Log_10_pg/ml).
(C) Viral load of non-SARS-CoV-2 viruses vs. age. Shading represents 95% CI for regression line slope. r^2^ and p-value for correlation are shown.

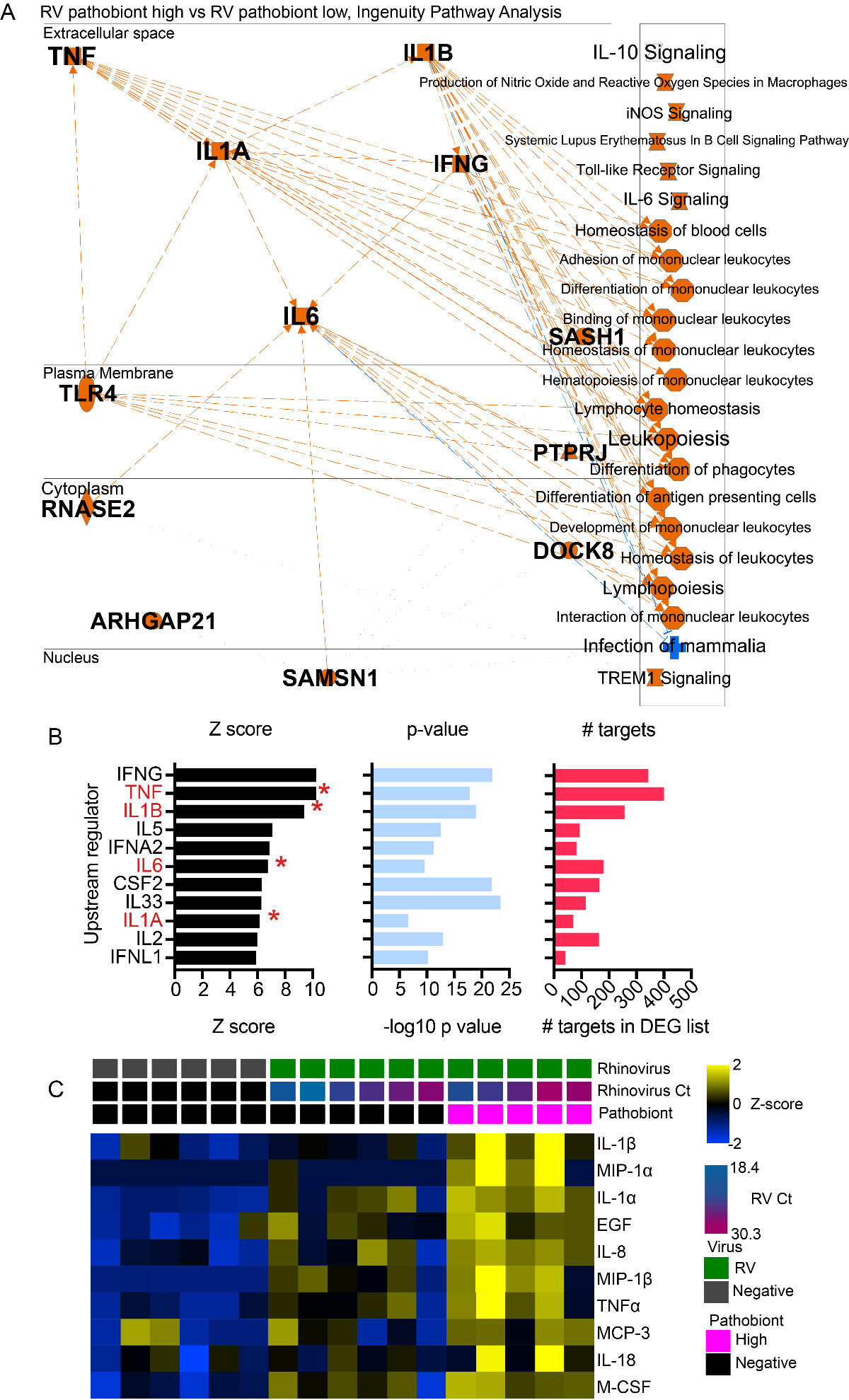

**Figure S2: Cytokines differentially expressed between rhinovirus-positive/pathobiont high vs. rhinovirus-positive/pathobiont low nasopharyngeal samples**

(A, B) Cytokine regulators of differentially expressed genes in RV-pathobiont high vs. RV-pathobiont low nasopharyngeal samples^1^. Upstream regulators are filtered on cytokines and listed in order of Z-score. Red asterisks denote cytokines which were also differentially expressed transcripts.

(C) Top differentially-expressed, upregulated cytokines in RV-bacterial high vs. RV bacteria low samples based on a multiplex immunoassay for 71 cytokines. Top cytokines differentiating RV from RV/pathobiont codetection are identified using two-group comparisons and are listed as the top 10 proteins in the order of most significant q-value.

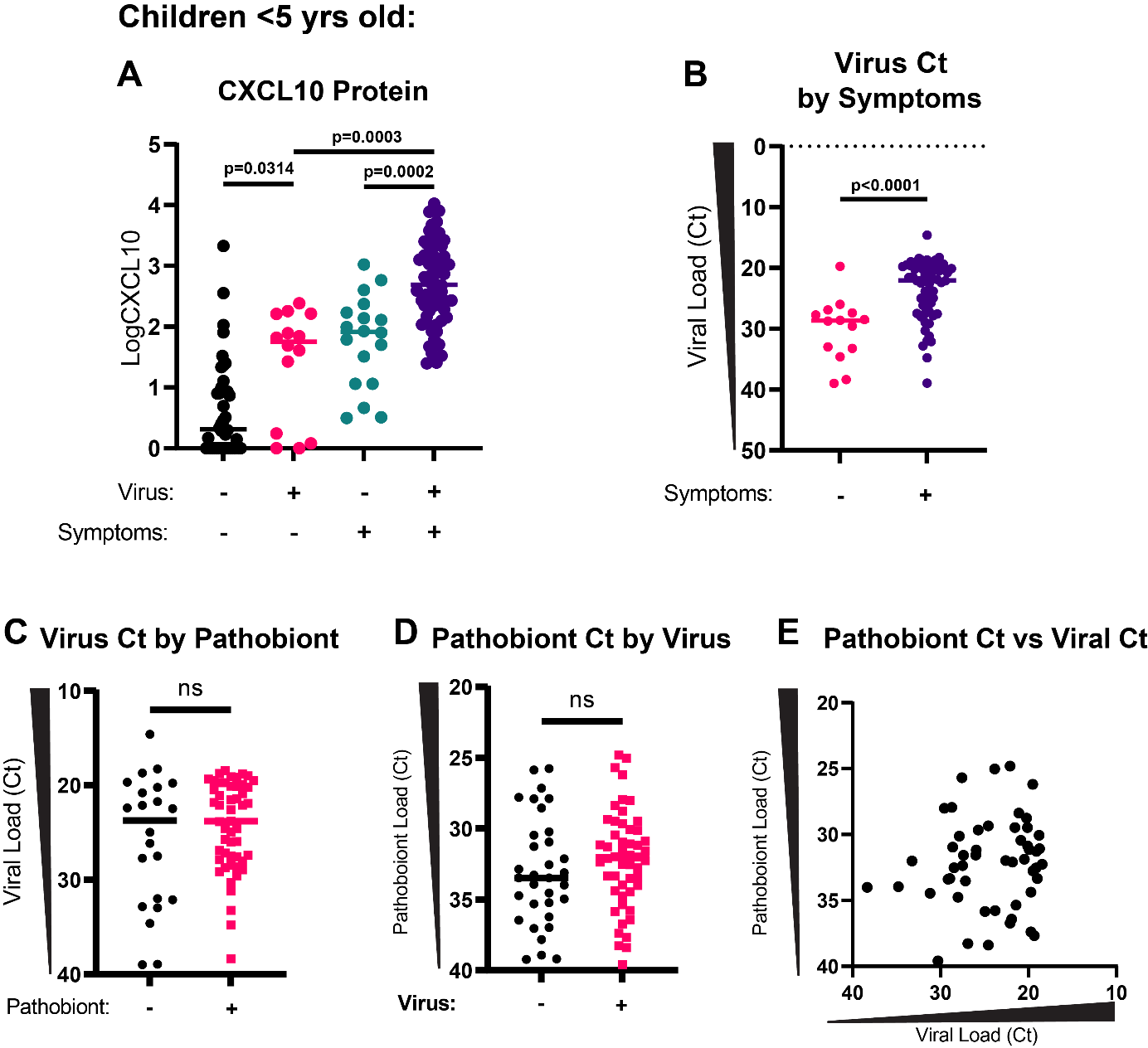

**Figure S3: Nasopharyngeal CXCL10 levels and viral loads based on symptom presentation and pathobiont status.**

(A) CXCL10 (Log_10_pg/mL) is shown for samples from <5-year-old children based on virus status and presence of respiratory virus-related symptoms. Statistical comparisons shown are performed with Welch’s one-way ANOVA test with Dunnett’s T3 multiple comparisons tests added.
(B) Viral load in virus-positive samples for <5-year-old children (n=73) based on presence of respiratory virus-related symptoms. Statistical comparison shown is performed with Welch’s t-test.
(C) Virus Ct value in virus-positive samples in <5-year-old children (n=73) in relation to pathobiont presence. Statistical comparison shown is performed with Welch’s t-test (p=0.3451).
(D) Pathobiont Ct value in pathobiont-positive samples for <5-year-old children (n=84) in relation to virus presence. Statistical comparison shown is performed with an unpaired t-test (p=0.2950).
(E) Pathobiont Ct is plotted against virus Ct in pathobiont/virus positive samples (n=51). Linear regression is not significant (p=0.4076).

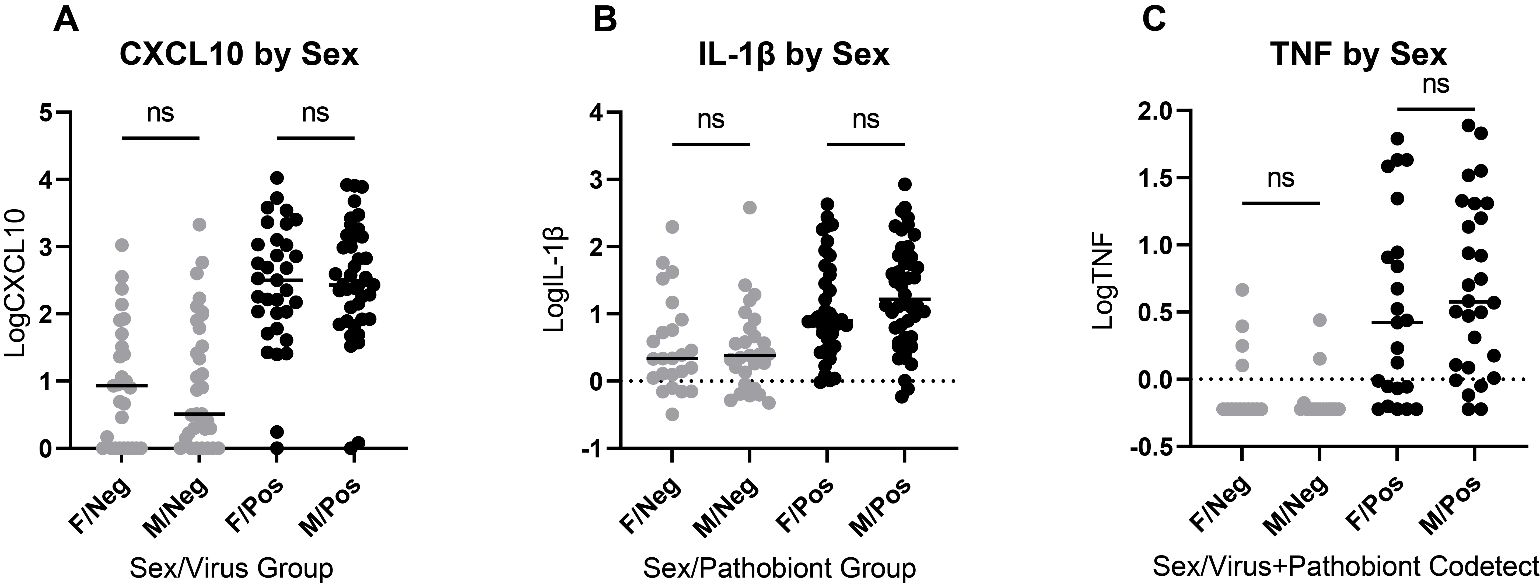

**Figure S4 (related to Figure 5): Mucosal cytokines in relation to virus and/or pathobiont positivity and biological sex.**

(A) In all samples tested for CXCL10 (n=135, <5 yrs), CXCL10 is shown according to sex and viral infection. Kruskal-Wallis test with Dunn’s multiple comparisons test is used. ns=not significant.
(B) In all samples tested for IL-1β (n=135, <5 yrs), IL-1β is shown according to sex and viral infection. Kruskal-Wallis test with Dunn’s multiple comparisons test is used. ns=not significant.
(C) In all samples tested for TNF (n=135, <5 yrs), TNF is shown according to sex and pathobiont/virus codetection. Kruskal-Wallis test with Dunn’s multiple comparisons test is used. ns=not significant.

| **Viral Infections** | | | **Pathobiont Detections** | | |
| --- | --- | --- | --- | --- | --- |
| **Result** | **0-19 yrs** | **<5 yrs** | **Result** | **0-19 yrs** | **<5 yrs** |
| **Negative** | **185** | **62** | **Negative** | **183** | **51** |
| **Single Infections** | **96** | **67** | **Single Detections** | **78** | **55** |
| SARS-CoV-2 | 57 | 36 | *M. catarrhalis* | 65 | 47 |
| RV | 10 | 9 | *S. pneumoniae* | 8 | 7 |
| Adeno | 8 | 7 | *H. influenzae* | 5 | 1 |
| hMPV | 8 | 4 |  |  |  |
| RSV | 4 | 3 |  |  |  |
| CoV 229E/OC43 | 3 | 2 |  |  |  |
| PIV 123 | 3 | 3 |  |  |  |
| CoV NL63/HKU1 | 2 | 2 |  |  |  |
| Flu A | 1 | 1 |  |  |  |
| Flu B | 0 | 0 |  |  |  |
| PIV 4 | 0 | 0 |  |  |  |
| **Co-infections** | **10** | **6** | **Co-detections** | **29** | **29** |
| SARS-CoV-2-Flu A | 3 | 0 | *M. cat/S.pneumo* | 16 | 16 |
| SARS-CoV-2-RV | 2 | 2 | *M.cat/H.inf* | 8 | 8 |
| SARS-CoV-2-229E/OC43 | 1 | 1 | *M.cat/S.pneumo/H.inf* | 3 | 3 |
| SARS-CoV-2-RSV | 1 | 1 | *H.inf/S.pneumo* | 2 | 2 |
| SARS-CoV-2/hMPV | 1 | 0 |  |  |  |
| hMPV-Adeno | 1 | 1 |  |  |  |
| hMPV-229E/OC43 | 1 | 1 |  |  |  |
| **Total** | **291** | **135** | **Total** | **290** | **135** |

**Table S1 (related to Figure 2, Table 1): Number of samples testing positive for respiratory viruses or bacterial pathobionts.**

|  | **Overall** | **Pathobiont Negative** | **Pathobiont Positive** | **p-value** |
| --- | --- | --- | --- | --- |
| N | 290 | 183 | 107 |  |
| **Sex** |  |  |  | 0.095 |
| Male | 144 | 84 | 60 |  |
| Female | 146 | 99 | 47 |  |
| **Median Age** | **5.85** | **11.4** | **2.2** | **<0.0001** |
| **Presentation (all samples)** |  |  |  | **<0.0001** |
| Symptomatic | 107 | 45 | 65 |  |
| Asymptomatic | 183 | 138 | 35 |  |
| **Presentation (all virus-positive, <5 yrs)** |  |  |  | 0.887078 |
| Symptomatic | 59 | 18 | 41 |  |
| Asymptomatic | 14 | 4 | 10 |  |
| **Viral Status** |  |  |  | **<0.0001 (all virus-positive)**  **0.000945 (SARS-CoV-2)** |
| Negative | 185 | 136 | 49 |  |
| Positive | 106 | 47 | 58 |  |
| SARS-CoV-2 only | 56 | 28 | 28 |  |

**Table S2 (related to Figure 2, Table 1): Overview of pathobiont test results by demographics, presentation, and viral status.**

Pathobiont test results are shown with respect to biological sex, age, presentation groups, and viral status. Chi-square tests are used for biological sex, presentation groups, and viral status. The Mann-Whitney test is used for pathobiont test result by age.
